# Perceptions and behavioural responses of the general public during the COVID-19 pandemic: A cross-sectional survey of UK Adults

**DOI:** 10.1101/2020.04.01.20050039

**Authors:** Christina J Atchison, Leigh Bowman, Charlotte Vrinten, Rozlyn Redd, Philippa Pristerà, Jeffrey W Eaton, Helen Ward

## Abstract

**Objective:** To examine risk perceptions and behavioural responses of the UK adult population during the early phase of the COVID-19 epidemic in the UK.

**Design:** A cross-sectional survey

**Setting:** Conducted with a nationally representative sample of UK adults within 48 hours of the UK Government advising the public to stop non-essential contact with others and all unnecessary travel.

**Participants:** 2,108 adults living in the UK aged 18 years and over. Data were collected between March 17 and 18 2020.

**Main outcome measures:** Descriptive statistics for all survey questions, including the number of respondents and the weighted percentages. Logistic regression was used to identify sociodemographic variation in: (1) adoption of social-distancing measures, (2) ability to work from home, and (3) willingness and (4) ability to self-isolate.

**Results:** Overall, 1,992 (94.2%) respondents reported taking at least one preventive measure: 85.8% washed their hands with soap more frequently; 56.5% avoided crowded areas and 54.5% avoided social events. Adoption of social-distancing measures was higher in those aged over 70 compared to younger adults aged 18 to 34 years (aOR:1.9; 95% CI:1.1 to 3.4). Those with the lowest household income were six times less likely to be able to work from home (aOR:0.16; 95% CI:0.09 to 0.26) and three times less likely to be able to self-isolate (aOR:0.31; 95% CI:0.16 to 0.58). Ability to self-isolate was also lower in black and minority ethnic groups (aOR:0.47; 95% CI:0.27 to 0.82). Willingness to self-isolate was high across all respondents.

**Conclusions:** The ability to adopt and comply with certain NPIs is lower in the most economically disadvantaged in society. Governments must implement appropriate social and economic policies to mitigate this. By incorporating these differences in NPIs among socio-economic subpopulations into mathematical models of COVID-19 transmission dynamics, our modelling of epidemic outcomes and response to COVID-19 can be improved.

## INTRODUCTION

On 31 December 2019, Chinese authorities notified the World Health Organisation (WHO) of an outbreak of pneumonia in Wuhan City, which was later classified as a new disease: COVID-19 (1). Following identification of cases in countries outside China, on 30 January 2020, WHO declared the outbreak of COVID-19 a “Public Health Emergency of International Concern” (1). In the UK, the first cases of COVID-19 were diagnosed at the end of January 2020, and community transmission was reported a few weeks later (2, 3). Government measures to control the epidemic were first announced on 22 January 2020 and included travel advice, information for those returning from affected countries, testing of suspected cases, isolation and contact tracing. This was followed in early February by a public health information campaign advising people to adopt hygiene measures to protect themselves and others, including more frequent handwashing with soap and water, using hand sanitiser if soap and water are not available, and covering mouth and nose with a tissue or sleeve when coughing or sneezing (4). Then, on 3 March 2020, the UK Government published its action plan setting out the UK-wide response to the novel coronavirus. The UK Government’s response outlined measures in four key areas: containing the outbreak, delaying its spread, mitigating the impact and research to improve diagnostics and treatment (5).

On March 16 2020, five days after the WHO declared the outbreak of COVID-19 a pandemic, the UK Prime Minister announced a shift to the delay phase of the UK response with measures aimed at suppressing the spread of the infection in the population through non-pharmaceutical interventions (NPIs), including social distancing of the whole population, isolation of cases for 7 days and quarantine of their household members for 14 days (6). The public was advised to stop non-essential contact with others and all unnecessary travel: including working from home where possible and avoiding pubs, theatres, restaurants and other social venues (6). This shift in response was prompted by a mathematical modelling study which showed that a combination of social distancing of the entire population, home isolation of cases and household quarantine of their family members (and possible school and university closure) was required to suppress transmission to a level that would enable the NHS to cope with the surge in cases requiring hospital admission and ventilation (7).

The effect of NPIs to reduce transmission rates is dependent on compliance with public health advice on social distancing. In the initial stages of the UK epidemic, this advice was voluntary, and not enforced by the government. This was criticised due to concern that measures may not have the desired impact if a significant proportion of the population were unable or unwilling to comply. As such, this study aimed to assess reported behaviour and intention to comply with the NPIs, as recommended by the UK Government at the time of the survey. Preliminary findings were shared with the Scientific Advisory Group for Emergencies (SAGE), which advises the UK Government’s response to COVID-19 (8).

## METHODS

### Study design and sample

A cross-sectional survey of a nationally representative sample of the UK adult (aged 18 years and over) population was conducted between March 17-18^th^ 2020, which followed the UK Government’s 16 March announcement to increase social distancing measures by advising the public to stop non-essential contact with others and all unnecessary travel.

The online survey was administered by YouGov, a market research company, to members of its UK panel of 800,000+ individuals as part of their omnibus survey (9). A sample of 2,108 adults was achieved through YouGov’s non-probabilistic active sampling method (9). Emails were sent to panellists from the base sample, randomly selecting panellists with particular characteristics to achieve quotas that matched the proportions of people with those characteristics in the UK 2011 census data (10). The responding sample was weighted to be representative of the UK adult population.

### Survey Instrument

The questionnaire was adapted from a survey used in a similar study conducted in Hong Kong (11). The questionnaire had four components: (1) socio-demographic characteristics, (2) risk perceptions towards COVID-19, (3) preventive behaviours, and (4) willingness and ability to self-isolate.

*Socio-demographic characteristics* consisted of sex, age, ethnicity, marital status, caring responsibilities, UK area of residence, and socio-economic status (SES). SES was assessed using five indicators: education level, employment status, household income, savings, and household tenure.

*Risk perceptions towards COVID-19* were measured by perceived susceptibility and perceived severity. Susceptibility was measured by asking respondents about perceived likelihood of being infected with COVID-19 under the UK Government’s current preventive measures. Severity was measured by asking respondents about perceived seriousness of symptoms if they were infected with COVID-19.

*Preventive behaviours* included information on perceived effectiveness and actual adoption of preventive behaviours (to protect oneself and others), to prevent both contracting COVID-19 and onward transmission, and were collected under three categories: (1) hygiene practices (wearing a face mask, washing hands more frequently with soap and water, using hand sanitiser more regularly, disinfecting the home, covering nose and mouth when sneezing or coughing) (2) travel avoidance (travel to affected countries and travel to areas inside and outside the UK, regardless of whether they were affected) (3) social distancing (avoiding public transport, social events, going out in general, going to hospital or other healthcare settings, crowded places, and contact with people who have a fever or respiratory symptoms).

*Willingness and ability to self-isolate* if asked by a healthcare professional were measured using two questions developed for this survey. At the time the survey was conducted, Public Health England’s operational definition of ‘self-isolation’ was “*if you have symptoms of coronavirus infection* (*COVID-19*), *however mild, do not leave your home* (*even to buy food or essentials*) *or have any visitors for 7 days from when your symptoms started. This includes not going to work, school or other public places, and avoiding public transport or taxis. Self-isolation is the same as voluntary quarantine*.”(12)

We worked with YouGov to optimise question clarity and ease of understanding for the UK population.

The survey instrument is freely available to download from the School of Public Health, Imperial College London COVID-19 resources webpage: https://www.imperial.ac.uk/mrc-global-infectious-disease-analysis/covid-19/covid-19-resources/

### Data Collection

Data were collected between 1630 GMT on 17 March 2020 and 1030 GMT on 18 March 2020. Participants identified for the sample were sent an email with a survey link. YouGov returned the anonymised data set to the Imperial College London research team for analysis.

### Data Analysis

Analyses were conducted in Stata 15 and SPSS version 25.

Descriptive statistics for all variables present the number of respondents and the weighted percentages.

Logistic regression was used to identify sociodemographic variation in: (1) adoption of social-distancing measures, (2) ability to work from home, and (3) willingness and (4) ability to self-isolate. Adoption of social distancing measures was proxied by respondents reporting to have avoided crowded places and social events to protect themselves or others from COVID-19.

Variables that appeared to be associated (p<0.20) in the unadjusted analyses were considered in the adjusted analyses. Adjusted odds ratios (aOR) and 95% confidence intervals (CI) were estimated. Associations with a p-value <0.05 in the adjusted analyses were considered to be statistically significant.

We tested for collinearity between education level, employment status, household income, savings and household tenure. For these categorical variables, collinearity was measured by examining bivariate relationships using Pearson’s Chi-squared tests. Where collinearity was detected we ran separate adjusted regression analyses for those variables, using only other explanatory variables in those models that were not strongly correlated.

### Patient and Public Involvement

We distributed an online feedback form to communities across the UK using local networks of public partners and contacts, Twitter and via VOICE-global.org, an online platform for public involvement in research established by Newcastle University. We received 420 responses, including 328 from members of the public. The experiences and feedback shared guided our study design and scope, including the phrasing of the survey tool’s closed-ended questions and the refinement of pre-populated answer choices.

Study results will be shared with the public both by posting on the VOICE-global.org news page, on the research team’s website and through direct mail with those who consented to be contacted about our research and involvement activity.

## RESULTS

The overall sample was designed to be representative of the UK adult population and is described in Table 1. In summary, of the 2,108 respondents, 11.1% were 18 to 24 years old, and 13.5% were 70 years or older. The majority of respondents were white (93.9%). In total, 43.4% were in full-time work and 14.1% were in part-time work.

**Table 1.**
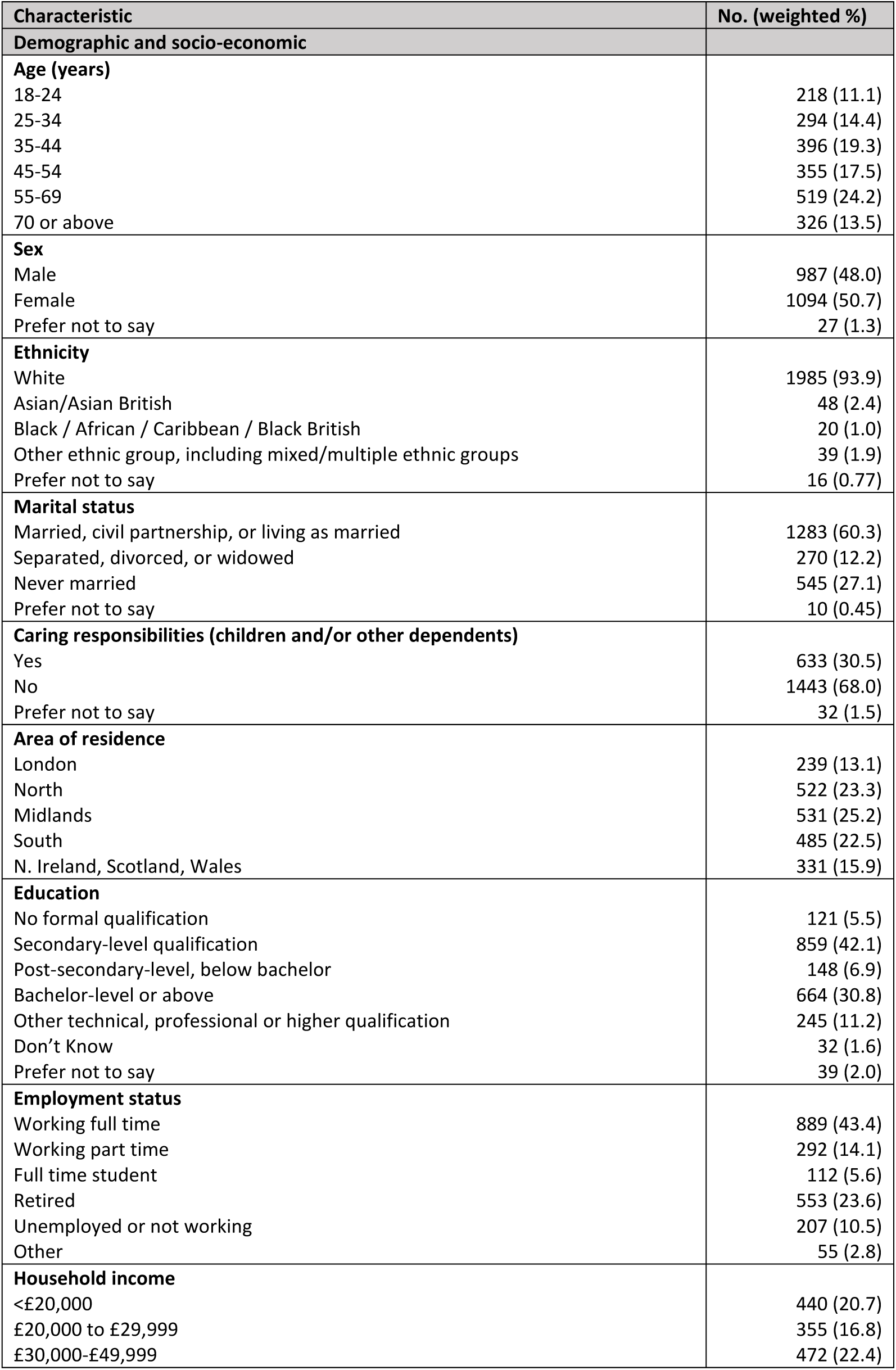
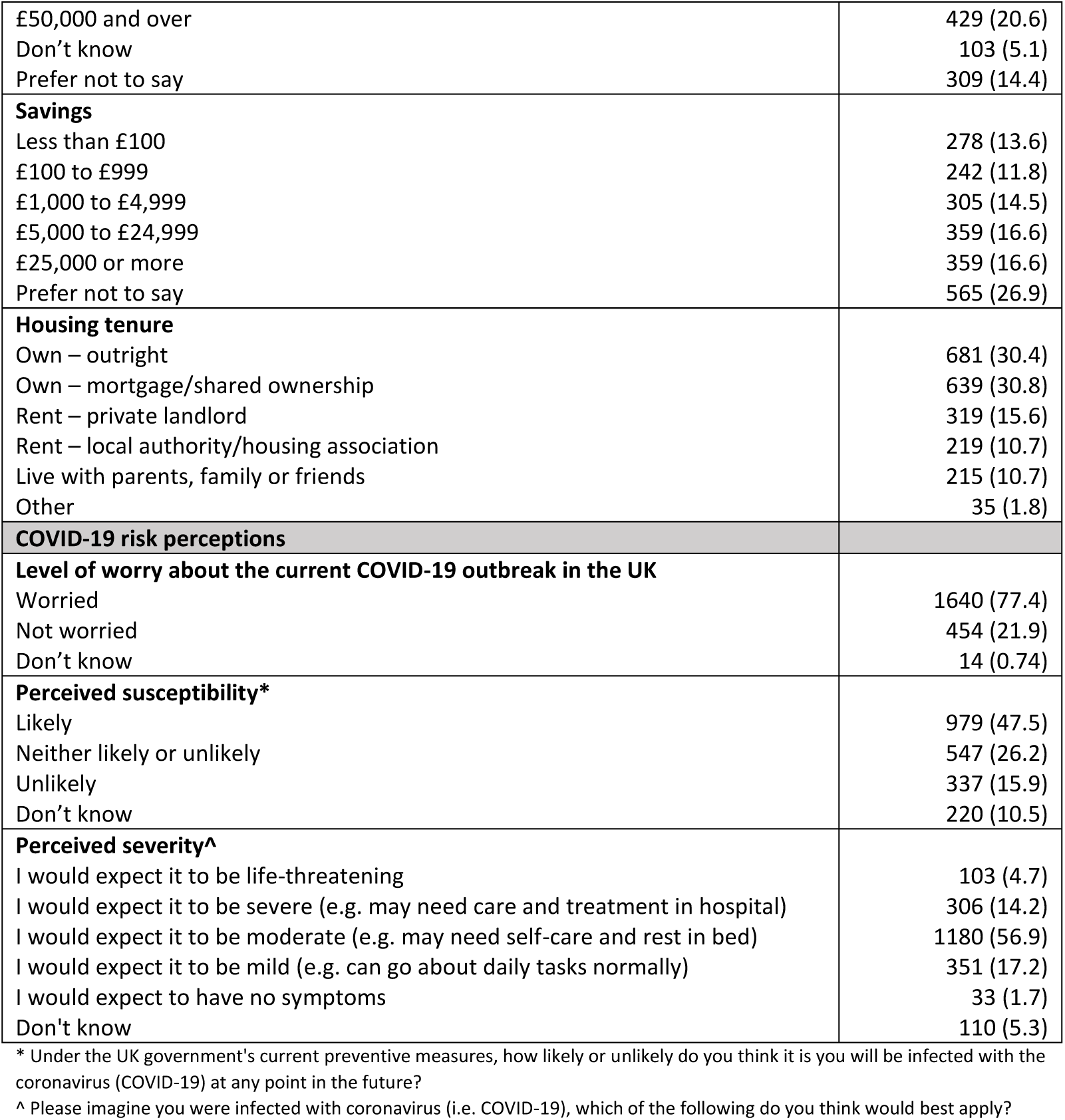
Demographics, socio-economic characteristics and COVID-19 risk perceptions, N=2,108

Overall, 77.4% (1,640/2,108) of respondents reported being worried about the COVID-19 outbreak in the UK. For those that had not previously tested positive for COVID-19, 47.5% (979/2,108) believed that it was likely they would be infected at some point in the future under the UK Government’s preventive measures. If infected, just over half (56.9%) of respondents would expect to be moderately severely affected (e.g. may need self-care and rest in bed) (Table 1).

Accordingly, 94.2% of adults reported taking at least one preventive measure (to protect oneself and others) against COVID-19 infection: 85.8% washed their hands with soap more frequently; 56.5% avoided crowded areas; 54.5% avoided social events and 39.2% avoided public transport (Figure 1). Most reported that their behaviour change was in response to government guidance (71.3%). Preventive measures perceived to be most effective were washing hands more frequently with soap and water (92.5%), avoiding contact with people who have a fever or respiratory symptoms (91.4%), and covering nose and mouth when sneezing or coughing (90.0%) (Figure 1). Perceived effectiveness of preventive measures was higher than actual adoption for all measures. This was particularly marked for social distancing measures (Figure 1).

**Figure 1.**
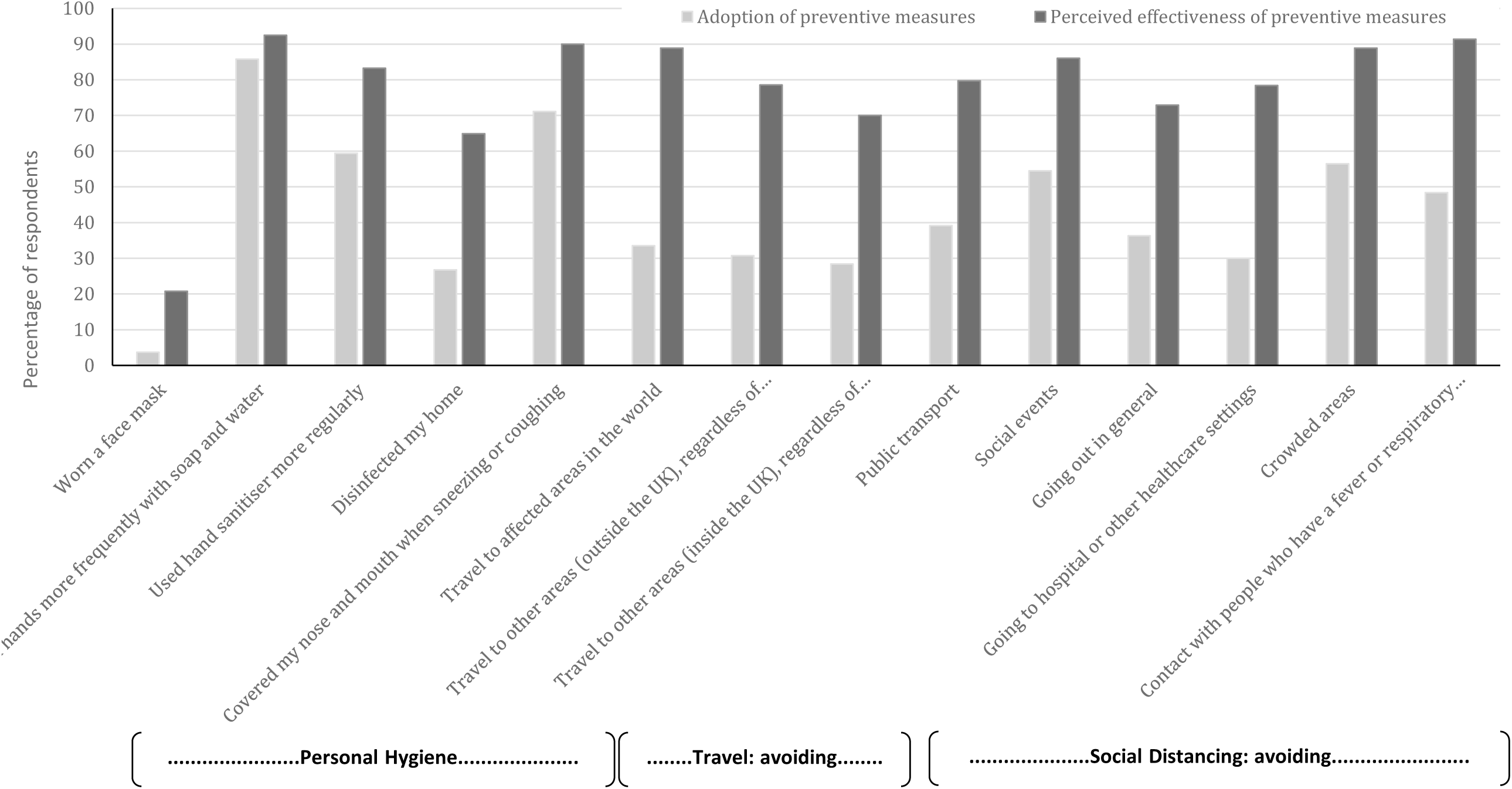
Perceived effectiveness and actual adoption of preventative measures to prevent transmission of COVID-19; N=2,108.

### Adoption of social-distancing measures

Overall, 45.2% of respondents reported adopting social distancing measures (avoiding crowded places and avoiding social events) to protect themselves or others from COVID-19.

Table 2 shows the regression analysis results for adoption of social-distancing measures. Being 70 years or older (64.2% vs. 38.4%; aOR:1.9; 95% CI:1.1,3.4) was positively associated with greater adoption compared to younger adults aged 18 to 34 years. Compared with those who were married, in a civil partnership, or living as married (48.4%), respondents who were separated, divorced, or widowed (44.1%; aOR:0.63; 95% CI:0.43,0.91) or never married (38.4; aOR:0.70; 95% CI:0.50,0.97) were less likely to have adopted social distancing measures to prevent transmission of COVID-19.

**Table 2.** Social distancing behaviour and ability to work from home by a range of sociodemographic factors, N=2,108

|  | Social distancing measures being taken yes vs no <sup>a</sup> |  |  | Able to work from home yes vs no (N=1,149) <sup>d</sup> |  |  |
| --- | --- | --- | --- | --- | --- | --- |
|  | N<br>(weighted %) | Unadjusted OR<br>(95% CI) | Adjusted OR<br>(95% CI) | N<br>(weighted %) | Unadjusted OR<br>(95% CI) | Adjusted OR<br>(95% CI) |
| **Social distancing measures being taken** |  |  |  |  |  |  |
| Yes | 969 (45.2) |  |  |  |  |  |
| No | 1,139 (54.8) |  |  |  |  |  |
| **Ability to work from home** |  |  |  |  |  |  |
| Yes |  |  |  | 540 (44.3) |  |  |
| No |  |  |  | 609 (53.0) |  |  |
| Don't Know |  |  |  | 32 (2.7) |  |  |
| **Age** |  |  |  |  |  |  |
| 18-34 | 202 (38.4) | Ref | Ref<sup>b</sup> | 154 (48.0) | Ref | Ref<sup>e</sup> |
| 35-49 | 229 (40.8) | 1.1 (0.86-1.4) | 1.2 (0.83-1.7) | 220 (48.0) | 1.0 (0.75-1.3) | 0.95 (0.63-1.4) |
| 50-69 | 329 (46.4) | **1.4 (1.1-1.8)\*\*** | 1.2 (0.85-1.8) | 151 (39.9) | **0.72 (0.53-0.98)\*** | 0.68 (0.43-1.1) |
| 70 or older | 209 (64.2) | **2.9 (2.2-3.8)\*\*\*** | **1.9 (1.1-3.4)\*** | 15 (53.9) | 1.3 (0.57-2.8) | 2.0 (0.57-7.0) |
| **Sex** |  |  |  |  |  |  |
| Male | 436 (42.9) | Ref | Ref<sup>b</sup> | 280 (46.1) | Ref | - |
| Female | 519 (47.4) | **1.2 (1.0-1.4)\*** | 1.1 (0.90-1.4) | 254 (44.8) | 0.95 (0.75-1.2) |  |
| **Ethnicity** |  |  |  |  |  |  |
| White | 919 (45.5) | Ref | - | 506 (45.1) | Ref | Ref<sup>e</sup> |
| BAME | 45 (42.1) | 0.87 (0.58-1.3) |  | 31 (54.2) | 1.4 (0.84-2.5) | 1.2 (0.56-2.4) |
| **Marital status** |  |  |  |  |  |  |
| Married, civil partnership, or living as married | 628 (48.4) | Ref | Ref<sup>b</sup> | 366 (47.2) | Ref | - |
| Separated, divorced, or widowed | 121 (44.1) | 0.84 (0.64-1.1) | **0.63 (0.43-0.91)\*** | 43 (39.5) | 0.73 (0.48-1.1) |  |
| Never married | 214 (38.4) | **0.66 (0.54-0.82)\*\*\*** | **0.70 (0.50-0.97)\*** | 128 (43.5) | 0.86 (0.66-1.1) |  |
| **Caring responsibilities** |  |  |  |  |  |  |
| Yes | 288 (45.8) | Ref | - | 213 (44.7) | Ref | - |
| No | 671 (45.4) | 0.98 (0.81-1.2) |  | 324 (46.4) | 1.1 (0.84-1.4) |  |
| **Area of residence** |  |  |  |  |  |  |
| London | 111 (45.2) | Ref | - | 76 (54.0) | Ref | Ref<sup>e</sup> |
| North of England | 220 (41.6) | 0.86 (0.63-1.2) |  | 129 (44.7) | 0.69 (0.45-1.0) | 1.1 (0.63-1.8) |
| Midlands and East of England | 249 (46.0) | 1.0 (0.76-1.4) |  | 123 (44.4) | 0.68 (0.45-1.0) | 0.93 (0.54-1.6) |
| South of England | 221 (45.0) | 0.99 (0.72-1.4) |  | 151 (49.0) | 0.82 (0.54-1.2) | 1.1 (0.66-1.8) |
| Northern Ireland, Scotland, Wales | 168 (49.9) | 1.2 (0.86-1.7) |  | 61 (35.2) | **0.46 (0.29-0.74)\*\*\*** | 0.57 (0.31-1.0) |
| **Education** |  |  |  |  |  |  |
| Degree or above | 321 (48.0) | Ref | - | 289 (62.6) | Ref | Ref<sup>f</sup> |
| Post-secondary but below degree | 186 (46.7) | 0.95 (0.74-1.2) |  | 105 (47.7) | **0.54 (0.39-0.76)\*\*\*** | **0.58 (0.41-0.82)\*\*** |
| Secondary qualifications or below | 436 (43.7) | 0.84 (0.69-1.0) |  | 137 (29.4) | **0.25 (0.19-0.33)\*\*\*** | **0.29 (0.21-0.39)\*\*\*** |
| **Employment status** |  |  |  |  |  |  |
| Working full time (30 or more hrs/wk) | 344 (38.6) | Ref | Ref<sup>b</sup> | 439 (48.9) | Ref | Ref<sup>e</sup> |
| Working part time | 136 (46.8) | **1.4 (1.1-1.8)\*\*** | 1.2 (0.85-1.7) | 101 (35.0) | **0.56 (0.42-0.74)\*\*\*** | 0.74 (0.50-1.1) |
| Full time student | 43 (36.3) | 0.91 (0.60-1.4) | 1.4 (0.71-2.6) | N/A | N/A | N/A |
| Retired | 331 (59.7) | **2.4 (1.9-2.9)\*\*\*** | 1.5 (0.99-2.3) | N/A | N/A | N/A |
| Unemployed / Not working | 95 (45.4) | 1.3 (0.97-1.8) | 1.4 (0.90-2.2) | N/A | N/A | N/A |
| **Household income** |  |  |  |  |  |  |
| £50,000 and over | 178 (41.0) | Ref | Ref<sup>c</sup> | 241 (67.3) | Ref | Ref<sup>f</sup> |
| £30,000 to £49,999 | 211 (43.8) | 1.1 (0.86-1.5) | 1.0 (0.77-1.4) | 131 (42.6) | **0.36 (0.26-0.50)\*\*\*** | **0.36 (0.26-0.51)\*\*\*** |
| £20,000 to £29,999 | 173 (48.5) | **1.4 (1.0-1.8)\*** | 1.2 (0.87-1.6) | 64 (30.7) | **0.22 (0.15-0.31)\*\*\*** | **0.22 (0.15-0.32)\*\*\*** |
| <£20,000 | 218 (49.0) | **1.4 (1.1-1.8)\*** | 1.1 (0.79-1.5) | 31 (22.7) | **0.14 (0.09-0.23)\*\*\*** | **0.16 (0.09-0.26)\*\*\*** |
| **Savings** |  |  |  |  |  |  |
| £25,000 or more | 177 (48.4) | Ref | Ref<sup>c</sup> | 100 (59.9) | Ref | Ref<sup>f</sup> |
| £5,000 to £24,999 | 177 (48.7) | 1.0 (0.75-1.4) | 1.3 (0.94-1.8) | 131 (57.6) | 0.91 (0.60-1.4) | 0.77 (0.49-1.2) |
| £1,000 to £4,999 | 126 (40.7) | 0.73 (0.54-1.0) | 1.0 (0.72-1.4) | 89 (43.7) | **0.52 (0.34-0.80)\*\*** | **0.46 (0.29-0.72)\*\*** |
| £100 to £999 | 91 (37.4) | **0.64 (0.45-0.89)\*\*** | 0.89 (0.61-1.3) | 67 (40.1) | **0.45 (0.29-0.70)\*\*\*** | **0.41 (0.26-0.67)\*\*\*** |
| Less than £100 | 122 (43.5) | 0.82 (0.60-1.1) | 1.1 (0.79-1.6) | 54 (33.1) | **0.33 (0.21-0.52)\*\*\*** | **0.30 (0.18-0.49)\*\*\*** |
| **Housing tenure** |  |  |  |  |  |  |
| Own outright | 377 (55.0) | Ref | Ref<sup>c</sup> | 93 (41.0) | Ref | Ref<sup>f</sup> |
| Own with mortgage/shared ownership | 261 (40.8) | **0.56 (0.45-0.70)\*\*\*** | 0.87 (0.66-1.1) | 293 (55.0) | **1.8 (1.3- 2.4)\*** | 1.4 (0.96-2.1) |
| Rented from private landlord | 145 (45.0) | **0.67 (0.51-0.88)\*\*** | 1.1 (0.79-1.5) | 85 (39.8) | 0.95 (0.65-1.4) | 0.65 (0.40-1.0) |
| Rented from local authority/housing association | 94 (42.2) | **0.60 (0.44-0.82)\*\*** | 0.79 (0.56-1.1) | 16 (18.2) | **0.32 (0.17-0.59)\*\*\*** | **0.37 (0.18-0.75)\*\*** |
| Live with parents, family, or friends | 78 (35.4) | **0.45 (0.32-0.62)\*\*\*** | 0.84 (0.54-1.3) | 46 (43.6) | 1.1 (0.69-1.8) | 0.96 (0.54-1.7) |

<sup>a</sup>Those that report avoiding crowded areas AND social events; <sup>b</sup>Mutually adjusted for age, sex, marital status, employment status, household income, savings and housing tenure; <sup>c</sup>Adjusted for age, sex, marital status and employment status. <sup>d</sup>Excluding those who responded "Don't know"; <sup>e</sup>Mutually adjusted for age, ethnicity, UK area of residence, household income, savings and housing tenure; <sup>f</sup>Adjusted for age, ethnicity, UK area of residence and housing tenure; <sup>g</sup>Adjusted for age, ethnicity and UK area of residence. \*p<.05, \*\*p<.01, \*\*\*p<.001

### Ability to work from home

Overall, 44.3% of respondents reported being able to work from home (i.e. permitted by their employer and have the necessary equipment to do their job from home).

Respondents who held post-secondary but below degree-level (47.7%; aOR:0.58; 95% CI:0.41,0.82) and secondary or below level (29.4%; aOR:0.29; 95% CI:0.21,0.39) education qualifications were less likely to be able to work from home compared with those educated to degree level (62.6%) (Table 2). As with educational level, there was a household income and savings gradient with ability to work from home. Those with the lowest household income (<£20,000) were six times less likely to be able to work from home compared to those with household incomes of £50,000 and above (22.7% vs. 67.3%; aOR:0.16; 95% CI:0.09,0.26). Respondents with £100 savings or less were three times less likely to be able to work from home compared to those with £25,000 or more in savings (33.1% vs. 59.9%; aOR:0.33; 95% CI:0.21,0.52) (Table 2).

Compared with those who owned their home outright, those renting accommodation from a local authority or housing association were less likely to be able to work from home (18.2% vs. 41.0%; aOR:0.37; 95% CI:0.18,0.75).

### Willingness and ability to self-isolate

Overall, perceived ability (87.0%) and willingness (87.6%) to self-isolate for 7 days if asked by a healthcare professional were high.

In terms of socio-demographic associations, there was no effect of sex on perceived ability to self-isolate (Table 3). However, women were somewhat more willing to do so than men (94.9% vs. 91.8%; aOR:2.1; 95% CI:1.2,3.5). Respondents from ethnic minority backgrounds perceived themselves to be less able to self-isolate than respondents from White backgrounds (84.8% vs. 92.1%, aOR:0.47; 95% CI:0.27,0.82), although they were equally willing to do so (Table 3).

**Table 3.** Ability and willingness to self-isolate by sociodemographic factors

|  | Able to self-isolate yes vs no (N=2,002)<sup>a</sup> |  |  | Willing to self-isolate yes vs no (N=1,978)<sup>a</sup> |  |  |
| --- | --- | --- | --- | --- | --- | --- |
|  | N<br>(weighted %) | Unadjusted OR<br>(95% CI) | Adjusted OR<br>(95% CI) | N<br>(weighted %) | Unadjusted OR<br>(95% CI) | Adjusted OR<br>(95% CI) |
| **Self-isolation ability/willingness** |  |  |  |  |  |  |
| Yes, I would | 1,834 (87.0) |  |  | 1,847 (87.6) |  |  |
| No, I wouldn't | 168 (8.0) |  |  | 131 (6.2) |  |  |
| Don't know | 106 (5.0) |  |  | 130 (6.2) |  |  |
| **Age** |  |  |  |  |  |  |
| 18-34 | 466 (90.8) | Ref | Ref<sup>b</sup> | 457 (91.6) | Ref | Ref<sup>b</sup> |
| 35-49 | 494 (90.0) | 0.91 (0.60-1.36) | 1.26 (0.70-2.25) | 508 (94.2) | 1.53 (0.95-2.48) | 1.33 (0.65-2.74) |
| 50-69 | 614 (92.2) | 1.19 (0.79-1.80) | 1.51 (0.74-3.09) | 627 (93.6) | 1.35 (0.87-2.10) | 1.04 (0.46-2.34) |
| 70 or older | 262 (94.9) | **1.9 (1.03-3.56)\*** | 1.83 (0.52-6.43) | 255 (94.4) | 1.60 (0.87-2.94) | 1.47 (0.44-4.99) |
| **Sex** |  |  |  |  |  |  |
| Male | 878 (90.7) | Ref | Ref<sup>b</sup> | 878 (91.8) | Ref | Ref<sup>b</sup> |
| Female | 957 (92.5) | 1.26 (0.92-1.73) | 1.23 (0.78-1.94) | 969 (94.9) | **1.65 (1.15-2.37)\*\*** | **2.05 (1.22-3.45)\*\*** |
| **Ethnicity** |  |  |  |  |  |  |
| White | 1,737 (92.1) | Ref | Ref<sup>b</sup> | 1,751 (93.7) | Ref | Ref<sup>b</sup> |
| BAME | 89 (84.8) | **0.47 (0.27-0.82)\*\*** | **0.33 (0.15-0.72)\*\*** | 86 (87.8) | **0.48 (0.26-0.91)\*** | 0.70 (0.28-1.77) |
| **Marital status** |  |  |  |  |  |  |
| Married, civil partnership, or living as married | 1,128 (92.3) | Ref | Ref<sup>b</sup> | 1,143 (94.5) | Ref | Ref<sup>b</sup> |
| Separated, divorced, or widowed | 215 (90.7) | 0.80 (0.49-1.30) | 0.94 (0.44-2.02) | 219 (92.8) | 0.77 (0.44-1.35) | 0.74 (0.33-1.67) |
| Never married | 482 (90.3) | 0.77 (0.54-1.10) | 1.07 (0.60-1.91) | 477 (91.0) | **0.59 (0.40-0.87)\*\*** | 0.88 (0.45-1.73) |
| **Caring responsibilities** |  |  |  |  |  |  |
| Yes | 555 (90.2) | Ref | - | 573 (94.6) | Ref | - |
| No | 1,257 (92.3) | 1.29 (0.93-1.80) |  | 1,253 (93.0) | 0.76 (0.57-1.15) |  |
| **Area of residence** |  |  |  |  |  |  |
| London | 241 (91.6) | Ref | - | 243 (92.7) | Ref | - |
| North of England | 427 (91.2) | 0.93 (0.54-1.61) |  | 430 (93.7) | 1.17 (0.64-2.13) |  |
| Midlands and East of England | 465 (93.0) | 1.21 (0.69-2.12) |  | 465 (93.9) | 1.21 (0.67-2.20) |  |
| South of England | 408 (89.5) | 0.76 (0.45-1.30) |  | 411 (92.2) | 0.92 (0.51-1.64) |  |
| Northern Ireland, Scotland, Wales | 294 (92.7) | 1.16 (0.63-2.14) |  | 297 (94.3) | 1.29 (0.66-2.52) |  |
| **Education** |  |  |  |  |  |  |
| Degree or above | 584 (93.1) | Ref | Ref<sup>c</sup> | 591 (94.9) | Ref | Ref<sup>c</sup> |
| Post-secondary but below degree | 332 (90.2) | 0.68 (0.43-1.08) | **0.59 (0.36-0.97)\*** | 329 (90.9) | **0.53 (0.32-0.88)\*** | **0.50 (0.29-0.85)\*** |
| Secondary qualifications or below | 863 (91.3) | 0.78 (0.53-1.15) | 0.71 (0.47-1.09) | 873 (93.6) | 0.77 (0.50-1.21) | 0.99 (0.61-1.85) |
| **Employment status** |  |  |  |  |  |  |
| Working full time (30 or more hrs/wk) | 799 (91.4) | Ref | Ref<sup>b</sup> | 804 (93.6) | Ref | Ref<sup>b</sup> |
| Working part time | 255 (90.4) | 0.88 (0.55-1.39) | 0.85 (0.45-1.59) | 263 (92.9) | 0.90 (0.53-1.54) | 0.58 (0.29-1.16) |
| Full time student | 105 (93.8) | 1.37 (0.62-3.02) | 1.63 (0.50-5.38) | 97 (89.0) | 0.55 (0.29-1.06) | 0.49 (0.17-1.37) |
| Retired | 450 (95.1) | **1.84 (1.13-2.97)\*** | 1.22 (0.48-3.11) | 444 (94.7) | 1.21 (0.74- 1.96) | 0.78 (0.33-1.89) |
| Unemployed or not working | 185 (89.8) | 0.81 (0.49-1.34) | 1.06 (0.49-2.32) | 191 (94.1) | 1.06 (0.56-2.00) | 2.39 (0.75-7.54) |
| **Household income** |  |  |  |  |  |  |
| £50,000 and over | 405 (95.5) | Ref | Ref<sup>d</sup> | 393 (94.7) | Ref | Ref<sup>d</sup> |
| £30,000 to £49,999 | 417 (90.7) | **0.46 (0.27-0.81)\*\*** | **0.44 (0.25-0.77)\*\*** | 424 (93.6) | 0.81 (0.46-1.44) | 0.73 (0.40-1.33) |
| £20,000 to £29,999 | 311 (92.6) | 0.60 (0.33-1.12) | 0.54 (0.28-1.03) | 308 (91.9) | 0.63 (0.35-1.13) | 0.57 (0.30-1.06) |
| <£20,000 | 363 (88.3) | **0.36 (0.21-0.62)\*\*\*** | **0.31 (0.16-0.58)\*\*\*** | 383 (93.0) | 0.73 (0.41-1.30) | 0.71 (0.36-1.40) |
| **Savings** |  |  |  |  |  |  |
| £25,000 or more | 329 (95.6) | Ref | Ref<sup>d</sup> | 318 (94.1) | Ref | Ref<sup>d</sup> |
| £5,000 to £24,999 | 319 (95.2) | 0.93 (0.45-1.92) | 1.20 (0.56-2.59) | 317 (95.8) | 1.45 (0.72-2.92) | 1.48 (0.70-3.11) |
| £1,000 to £4,999 | 274 (92.3) | 0.55 (0.28-1.07) | 0.73 (0.36-1.48) | 272 (92.8) | 0.82 (0.44-1.55) | 0.83 (0.42-1.64) |
| £100 to £999 | 217 (90.0) | **0.42 (0.22-0.81)\*** | 0.60 (0.29-1.25) | 221 (94.8) | 1.18 (0.56-2.45) | 1.17 (0.53-2.60) |
| Less than £100 | 232 (84.4) | **0.25 (0.13-0.45)\*\*\*** | **0.35 (0.18-0.68)\*\*** | 250 (90.9) | 0.63 (0.34-1.16) | 0.55 (0.28-1.08) |
| **Housing tenure** |  |  |  |  |  |  |
| Own outright | 576 (93.8) | Ref | Ref<sup>d</sup> | 575 (95.0) | Ref | Ref<sup>d</sup> |
| Own with mortgage/shared ownership | 571 (92.1) | 0.77 (0.50-1.20) | 0.99 (0.58-1.68) | 584 (95.0) | 0.98 (0.59-1.64) | 0.87 (0.47-1.60) |
| Rented from private landlord | 277 (89.1) | **0.53 (0.33-0.86)\*** | 0.71 (0.39-1.29) | 297 (94.3) | 0.85 (0.47-1.55) | 0.82 (0.40-1.67) |
| Rented from local authority/housing association | 188 (87.9) | **0.49 (0.29-0.82)\*\*** | 0.68 (0.36-1.26) | 188 (90.0) | **0.47 (0.26-0.84)\*** | **0.39 (0.21-0.75)\*\*** |
| Live with parents, family, or friends | 197 (92.9) | 0.85 (0.46-1.56) | 1.34 (0.60-2.99) | 178 (88.1) | **0.39 (0.22-0.69)\*\*** | **0.43 (0.20-0.95)\*** |

<sup>a</sup>Excluding those who responded "Don't know"; <sup>b</sup>Mutually adjusted for age, gender, ethnicity, marital status, education, employment status, household income, savings, and housing tenure; <sup>c</sup>Adjusted for age gender, ethnicity, marital status, employment status and housing tenure; <sup>d</sup>Adjusted for age, gender, ethnicity, marital status, and employment status; \*p<.05, \*\*p<.01, \*\*\*p<.001

Some indicators of socioeconomic status were significantly associated with perceived ability and willingness to self-isolate. Respondents who held post-secondary but below degree-level education qualifications were less able (90.2% vs. 93.1%; aOR:0.59; 95% CI:0.36,0.97) and less willing (90.9% vs. 94.9% aOR:0.50; 95% CI:0.29,0.85) to self-isolate than respondents educated to degree level (Table 3). Those with household incomes below

£20,000 were three times less likely to be able to self-isolate compared with those on household incomes of £50,000 and above (88.3% vs. 95.5%; aOR:0.31; 95% CI:0.16,0.58). Similarly, respondents with less than £100 in savings were three times less likely to be able to self-isolate compared with those with savings of £25,000 or more (84.8% vs. 95.6%; aOR:0.35; 95% CI:0.18,0.68). There was no effect on willingness to self-isolate by household income or amount of savings (Table 3).

Those in accommodation rented from a private landlord, local authority, or housing association were less likely to report being able to self-isolate, although this association was no longer significant when other sociodemographic factors were adjusted for. In terms of willingness to self-isolate, respondents renting accommodation from a local authority or housing association (aOR:0.39; 95% CI:0.21,0.75) and those living with parents, family or friends (aOR:0.43; 95% CI:0.20,0.95) were less likely to be willing to self-isolate compared with those who owned their home outright (Table 3).

## DISCUSSION

This study reports on the perceptions and behaviour of the UK adult population in the two days following the government introduction of recommendations on social distancing (6). We found high levels of self-reported behaviour change. Notably, the most-adopted measures, washing hands more frequently with soap and water, using hand sanitiser and covering nose and mouth when sneezing or coughing, prominently featured in national public health campaigns from relatively early on in the epidemic (4) and mirrors results seen in previous pandemics (13). However, there were marked differences between the perceived effectiveness and adoption of NPIs. This suggests that lack of knowledge on what measures are effective against COVID-19 is not a key driver of compliance in the UK population. In contrast, a similar study conducted in Hong Kong showed comparatively high perceived effectiveness and adoption of preventive measures (11).

Our results highlighted significant differences across demographic and socio-economic strata for social distancing behaviour, ability to work from home, and the ability and willingness of people to self-isolate. Adoption of social distancing measures was almost twice as likely in people over 70 compared to adults aged 18 to 34 years. Notably, those that were single were less likely to practice social distancing. There was a strong association between socioeconomic deprivation and ability to adopt NPIs. Although willingness to self-isolate was high overall, those from more disadvantaged backgrounds were less likely to be able to work from home or self-isolate if needed, suggesting the existence of structural barriers to adopting preventive behaviours in these groups.

The strength of this study is in the representative sample of the UK adult population, the ability to achieve our sample size quickly, and the timeliness in relation to changing government recommendations. This study has three limitations. First, with an online approach, responses of those without internet access were under-represented. Second, the survey tool consisted of predominantly closed-ended questions. Thus, we were unable to explore responses in more depth. Third, surveys collecting self-report data are generally subject to limitations including honesty, introspective ability and interpretation of the questions.

Our findings highlight the stark choices faced by those in lower socio-economic groups and suggest that unless the government intervenes to support these individuals, the impact of this epidemic will likely be felt unequally in our society. Not only this, but if a large proportion of the population continues to work while unwell, low compliance will render the various forms of social distancing less effective, as low-income workers are forced to choose between financial and physical health. Indeed, this behaviour has already been observed in the workplace in previous pandemics: workers without access to paid sick leave were more likely to work while unwell than those with paid sick leave (14). A study in China after the H7N9 epidemic found that only 7% of people reported willingness to self-quarantine (15). Also, during the Middle East Respiratory Syndrome (MERS) outbreak in Korea in 2015, there was heterogeneous uptake of preventive interventions (16).

### Conclusions and policy implications

In the absence of a vaccine and treatments over the short-term, high compliance with social distancing, self-isolation and household quarantine is paramount to reduce transmission and the impact of COVID-19. And as the epidemic evolves, it is likely that compliance with preventive behaviours will continue to evolve too. NPI compliance, risk perception and behaviour are not consistent across cultures, social status or time. Indeed, previous studies have shown that perceptions and behaviours often change over time (13). Therefore, current modelling projections of the impact of NPIs on morbidity and mortality are always provisional (7). Future COVID-19 models should explore the variation captured in this and previous studies to better estimate the impact of differential uptake of NPIs in the UK and beyond. It is also important to monitor behaviour throughout the epidemic to know when to implement further public health messaging, and when further or alternative government actions might be required, to mitigate falling compliance.

Our findings highlight that those most economically disadvantaged in society are less able to comply with certain NPIs, likely in part due to their financial situation. Whilst one approach may be to better tailor public health messaging to this subpopulation, this must be done alongside considered fiscal and monetary policy to mitigate the financial costs of following government public health advice. Therefore, it is imperative that the UK Government, and governments around the world, quickly develop and implement policies to support the most vulnerable, in a bid to minimise the long-term social and economic harm caused by COVID-19. Government policy should recognise the disparity in impact across socio-economic groups, particularly across the labour market, and should aim to support workers equitably across the income spectrum. This would likely help increase compliance across the population to the levels required to suppress transmission and thereby reduce the strain on national health services, both in the UK and abroad. Although the UK Government has since announced a range of measures to support public services, individuals and businesses in part to facilitate compliance with current lockdown measures (17), it is uncertain how long these protections will be in place for and whether they will continue once lockdown restrictions are lifted.

In summary, the population’s response to public health advice is currently the key factor in tackling the COVID-19 epidemic and whether the curve is flattened sufficiently to allow health services to cope (7). Factors affecting uptake and compliance with preventive measures are critical. Those from socioeconomically more deprived backgrounds, in particular, may need further financial assurance and assistance from the government to be able to implement some of these measures, such as self-isolation.

## Data Availability

The survey instrument is freely available to download from the School of Public Health, Imperial College London COVID-19 resources webpage: https://www.imperial.ac.uk/mrc-global-infectious-disease-analysis/news--wuhan-coronavirus/covid-19-resources/
The data used for the analyses are publicly available from the corresponding author on request.

https://www.imperial.ac.uk/mrc-global-infectious-disease-analysis/news--wuhan-coronavirus/covid-19-resources/

## ACKNOWLEDGEMENTS

The survey was conducted by YouGov on behalf of the Patient Experience Research Centre, School of Public Health, Imperial College London. Questions were developed by the authors, and YouGov provided suggestions to improve clarity and understandability.

We thank Prof. Kin On Kwok, Ms. Wan In Wei, Prof. Samuel Yeung Shan Wong and the research team in the Division of Infectious Diseases of JC School of Public Health and Primary Care, The Chinese University of Hong Kong, Hong Kong Special Administrative Region, China for permission to use their survey instrument and translating it into English.

The study was supported by Imperial NIHR Research Capability Funding. Professor Ward is a National Institute for Health Research (NIHR) Senior Investigator and receives funding from The Imperial NIHR Biomedical Research Centre, the NIHR Applied Research Collaborative North West London and The Wellcome Trust. The views expressed in this article are those of the authors and not necessarily those of the NHS, the NIHR, or the Department of Health.

## CONTRIBUTORS

CJA, LB, RR, PP and HW designed the study. CJA, JWE and CV analysed the data and performed the statistical analyses. CJA, LB, RR, CV and HW drafted the initial manuscript. All authors reviewed the drafted manuscript for critical content and approved the final version. The corresponding author attests that all listed authors meet authorship criteria and that no others meeting the criteria have been omitted. CJA and HW are the guarantors.

## COMPETING INTERESTS

All authors have completed the ICMJE uniform disclosure form at www.icmje.org/coi_disclosure.pdf and declare: no financial relationships with any organisations that might have an interest in the submitted work in the previous three years; no other relationships or activities that could appear to have influenced the submitted work.

The lead authors (CJA and HW are the manuscript’s guarantors) affirm that the manuscript is an honest, accurate, and transparent account of the study being reported; that no important aspects of the study have been omitted; and any discrepancies have been explained.

## ETHICAL APPROVAL

The Imperial College London Research Ethics Committee approved the study (Ref 20IC5861).

Informed consent was obtained from those who chose to complete the survey after having read introductory information on its content and purpose.

## DATA SHARING

The survey instrument is freely available to download from the School of Public Health, Imperial College London COVID-19 resources webpage: https://www.imperial.ac.uk/mrc-global-infectious-disease-analysis/news--wuhan-coronavirus/covid-19-resources/

The data used for the analyses are publicly available from the corresponding author on request.

